# Extrafascial transfissural approach with finger fracture technique approach for liver resection.Old is still gold?

**DOI:** 10.1101/2020.04.08.20057562

**Authors:** Bhavin B Vasavada, Hardik Patel

**Affiliations:** Consultant Hepatobiliary and liver transplant surgeon, Shalby Hospitals, Ahmedabad

**Keywords:** Liver resection, Glissonian Pedical aprroach, Transfissural techniques, clamp crush technique

## Abstract

**Background:** We evaluated our protocol of extrafascial transfissural approach for liver resection with intrafascial approach that we use in case of donor hepatectomy.

**Material and Method:** We use extrafascial transfissural approach with finger fracture technique for liver resections and inftrafascial approach with clamp crush technique in case of donor hepatectomy. Major hepatectomy defined as resection of 2 or more adjacent segments.We compared these two techniques with regard to blood loss, operative time, morbidity and mortality.We also evaluated over all factors responsible for 90 days mortality.statistical analysis was done using SPSS version 23.(IBM).Categorical factors were evaluated using chi square test and numerical factors were analyzed using Mann Whitney U test. Multivariate analysis was done using logisitic regression method. Ethical approval for our clinical study was obtained by human research COA number SBI 3246.

**Results:** We evaluated 26 liver resections done in last three years. 19 liver resections were done using extrafascial transfissural approach for various liver tumors and 7 living donor hepatectomies were done using itrafascial technique with clamp crush methods. Mean age of patients was 50.73 years.16 patients were males and 10 were females. Mean blood loss was 273.9 ml and mean operative duration was 184.7 minutes. 22 were major resections, 4 were minor liver resections. All minor liver resections were in transfissural approach however there was no statistical significant difference between them. Being live liver doners patients in intrafacial group they were younger than extrafascial transfissural group. (p=0.01). There was no statistical significant difference in blood loss, blood products requirements, morbidity, in hospital and 90 days mortality in both the groups. However extrafascial transfissural with finger fracture technique was associated with significant less operative time. (168.13 minutes vs 222.86 minutes) (p=0.006). 90 days mortality was associated with higher ASA grade (0.018) and blood loss (0.008). However in multivariate analysis no factor indepedently predicted mortality.

**Conclusion:** Extrafascial transfissural approach significantly reduces operative time, without affecting morbidity and mortality in liver resection.

## Introduction

Couinaud was convinced that Glisson’s capsule was the most important component of the liver. Couinaud described three main approaches to the inflow system at the hepatic hilus; the intrafascial, the extrafascial, and the extrafascial and transfissural approach.^1,2^ The extrafascial approach was first described by Takasaki and Couinaud. The extrafascial approach constitutes an approach to the pedicles at the hepatic hilus without liver dissection.^1,2,3^ The extrafascial transfissural approach was first introduced by Tung and extrafascial transfissural approach with finger fracture by Lin.^4,5^

We compared our protocol of extrafascial transfissural approach with finger fracture techniques in liver resections with intrafascial approach with clamp crush technique in donor hepatectomies.

## Material and Methods

### Surgical Technique

#### Extrafascial Transfissural technique

We mobilize liver from attachments by dividing triangular ligaments, We start dividing main portal fissure for right and left hepatectomy, right portal fissure for right posterior secterectomy and left trisegmentectomy and left portal fissure for right trisegmentectomy and left lateral segmentectomy and loop glissonian pedical via extrafascial approach intrahepatically. We cut and ligate glissonian pedical en-mass. [Figure 1] and complete the liver transection.

**Figure 1:**
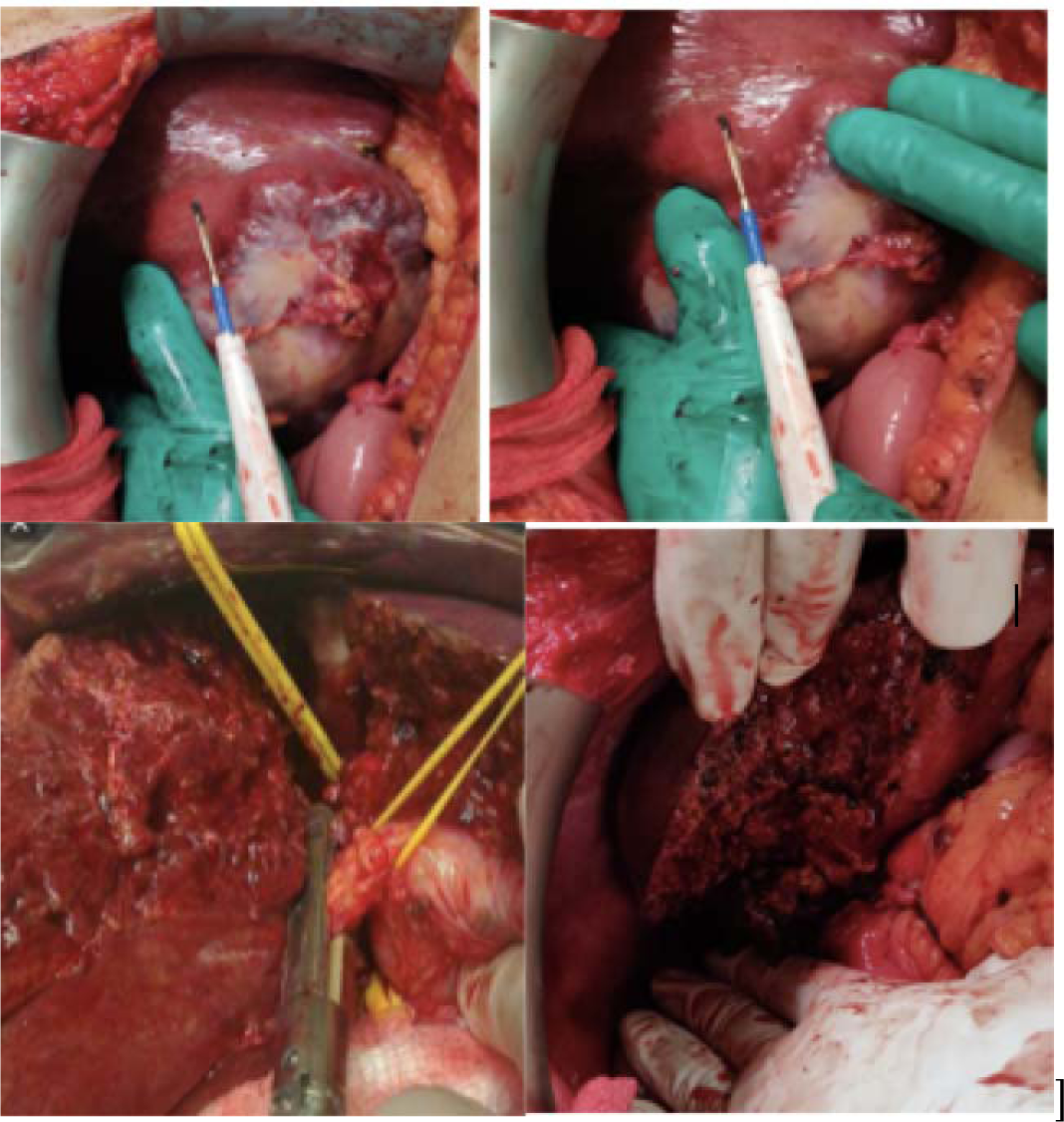
step of trasfissural extrafascial approach. 1,2) marking the main portal fissure.3) looping and enmass cutting of glissonian pedical with extrafascial approach.4) completed hepatectomy.

#### Intrafascial clamp crush technique in donor hepatectomy

All the hepatectomies in donor in this study were right hepatectomies. We loop right hepatic artery, right portal vein and right hepatic vein extrahepatically and then clamp right hepatic artery and right portal vein and mark the demarcation line. We transect liver by clamp crush technique. We take right lobe with or without middle hepatic vein based on graft to recipient weight ratio and liver remnant in donor. We take right lobe without middle hepatic vein if remnant is less than 32 percent.

Major hepatectomy defined as resection of 2 or more adjacent segments.We compared these two techniques with regard to blood loss, operative time, morbidity and mortality.

### Statistical analysis

Statistical analysis was done using SPSS version 23(IBM). Categorical factors were evaluated using chi square test and numerical factors were analyzed using Mann Whitney U test. Multivariate analysis was done using logisitic regression method. ROC curves were prepared for factors which are significantly different between extrafascial transfissural techniques and intrafascial clamp cursh techniques. P value less than 0.05 was considered as significant. Kaplan Meier survival curve prepared comparing survival in two different methods.

## Results

We performed twenty-six liver resection from February 2016 to February 2019. 16 liver resections were performed for various liver pathologies by using transfissural extrafascial approach with finger fracture techniques and 7 donor hepatectomies were performed using intrafascial approach using clamp crush technique. Mean age of entire cohort was 50.43. 13 patients were male and 10 were females. Mean blood loss was 273.9 ml, mean hospital stay was 4.8 days, mean ASA score was 2.3,mean operative time was 184.7 days. 19 were major hepatectomies and 4 were minor hepatectomies.

Trasfissural and extrafascial group and intrafascial group were compared. [Table 1].

**Table 1.** Comparisons between the extrafascial Transfissural group and the intrafascial group.

| Factors | ExtraFascial Transfissural<br>(n=19) | Intrafascial<br>(n=7) | P<br>value |
| --- | --- | --- | --- |
| Age(mean) (years) | 56.5 | 36.4 | 0.010 |
| Sex | Male= 11 ,Female= 8 | Males=5,female<br>=2 | 0.405 |
| Blood loss (ml) (Mean) | 268.7 | 285.7 | 0.154 |
| Operative time (min)<br>(Mean) | 168.1 | 222.8 | 0.027 |
| ASA Grade. (Mean) | 2.4 | 2.0 | 0.249 |
| Hospital stay. (Mean) | 4.6 | 5.2 | 0.033 |
| Major/minor<br>hepatectomy | Major=15, Minor=4 | Major=7, Minor=0 | 0.273 |
| 90 day mortality | 3 | 0 | 0.526 |
| Morbidity | 2 | 1 | 1.0 |

**Table 2.** Etiology for liver resections in the extrafascial transfissural approach with finger fracture technique.

| Etiology | Numbers |
| --- | --- |
| Hepatocellular Carcinoma | 6 |
| Carcinoma gall bladder | 8 |
| Cholangiocarcinoma | 3 |
| Colorectal Liver metastasis | 2 |

Age was significantly less in donor hepatectomies in the intrafascial group because younger and healthy donors were selected for donor hepatectomy. Both the groups were equal in terms of 90 days morbidity and mortality and ASA grade as well as major hepatectomies and blood loss.

However Operative time and Hospital stay were significantly less in extrafascial trransfissural group with finger fracture technique.

Kaplan Meier survival curve was prepared comparing two methods. However there was no statistical significant difference between two group of patients. 90 days mortality was associated with higher ASA grade (0.018) and blood loss (0.008). However in multivariate analysis no factor indepedently predicted mortality. [figure 2]

**Figure 2:**
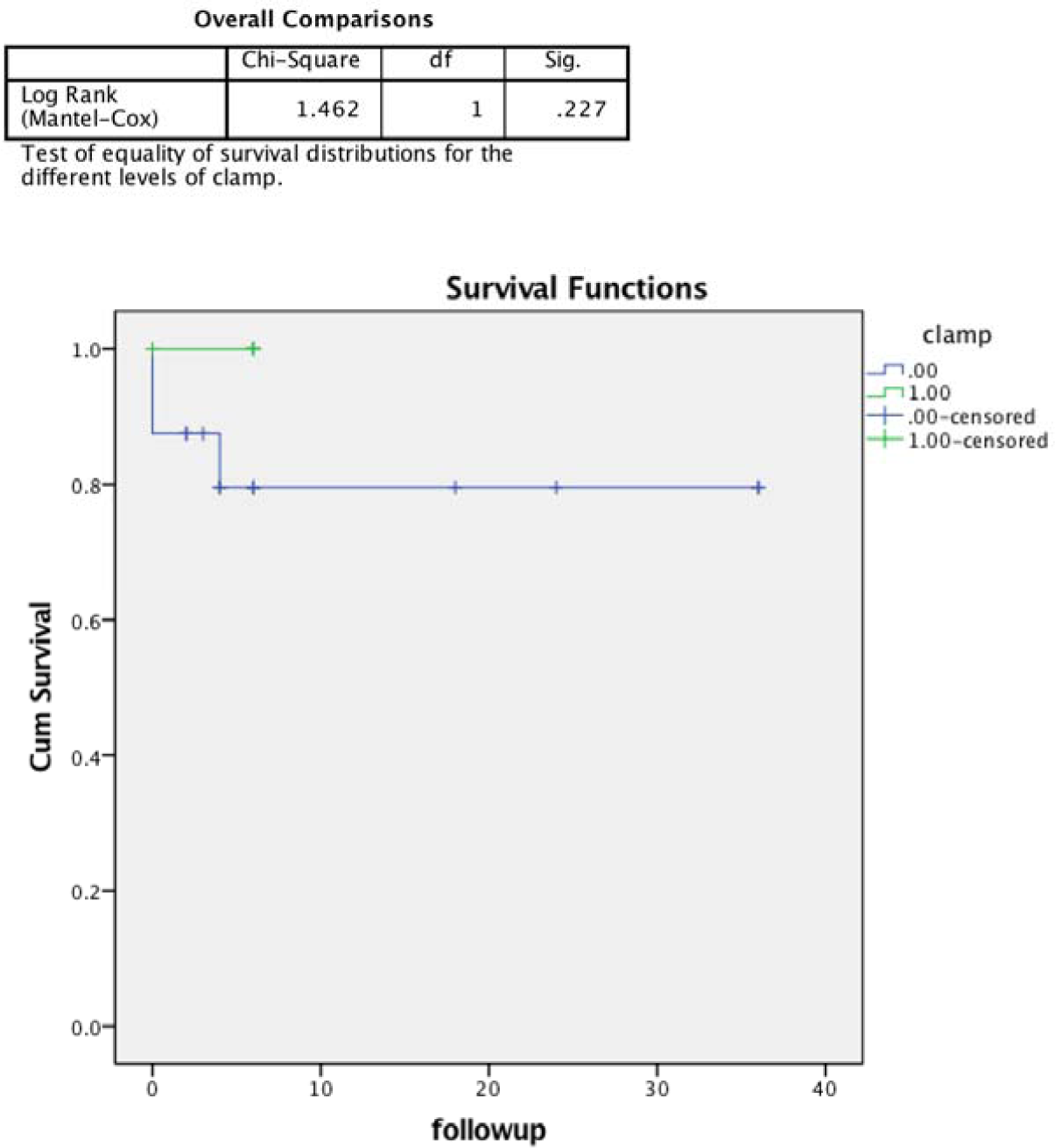
90 days Kaplan Meier survival curve between two groups of patients. There was no statistical significant difference between the groups. (p=0.227)

## Discussion

Liver resection techniques and outcomes have improved significantly in last 100 years. ^6^ Dr. Luis performed the first liver surgery but the patient died 6 hours later due to bleeding. The first successful liver resection is attributed to Dr. Langenbuch in 1888. ^7^

Many techniques of liver parenchyma trasection techniques have been described in literature like finger fracture,clamp crush, harmonic scalpel, ligasure, staplers, water jet. However none of the techniqes are superior to other techniques.^8^

Glissonian pedical approach popularized by takasaki et al^9^ and subsequently adopted by many surgeons due to its technical ease and same techniques are also adopted in laproscopic liver resection.^10,11,12^ Launois et al described posterior technique^13^, however we prefer antierior transfissural or intrahepatic approach as we feel it is less time consuming.

However, very few studied compared transfissural extrahepatic approach with glissonian pedicals with intrafascial clamp crush technique. We perofrmed donor hepatectomies via intrafascial approach and liver transection via clamp crush technique, rest all the liver resections were done using transfissural techniques. and liver transection was done using finger fracture techniques.In donor hepatectomy or intrafascial group patients were significantly younger as we chose healthy and young adults and liver donors. However extrafascial transfissural approach was associated with significantly less operative time and (p=0.027) and signifcantly less hospital stay (0.033). There was no significant difference between major and minor hepatectomies performed between the two groups.

There was no difference between morbidity and mortality between two groups. (p=1 and 0.5 respectiveluy) ans also there was no significant difference between blood loss in both the groups. (p=0.154) infact mean blood loss was less in extrafascial transfissural technique thout it did not reach statistical significance.Suggestiting extrafascial transfissural finger fracture technique significantly reduces operative time and hospital stay without increasing blood loss.

Mortality was associated with more blood loss and high ASA grade of patient (p=0.08 and p=0.018 respectively) however on multivariate analysis no factor indepedently predicted mortality. Virani et al ^14^ in their analysis also found ASA score as one of the factors associated with mortality. Dokmak et al also showed similar results.^15^ A study from mexico also showed blood loss as a major factor associated with mortality. ^16^ Kaplan Meier survival curve also showed no significant difference between the two groups. (p=0.227)

One patient developed post hepatectomy liver failure and one patient developed acute kidney injury. Majority of deaths (2) occurred in cholangiocarcinoma patients and with trisegmentectomies which co relates with findings in studies with large numbers studies.^17^

Being a retrospective study our study has inherent limitation with retrospective studies. Numbers of liver resections performed in our study is less and we are still in process of evaluting patients prospectively.Randomised control trial comparing these two techinies can give us more authentic analysis.

In conclusion transfissural extrahepatic approach with finger fracture technique decreases operative time and hospital stay without increasing blood loss, morbidity and 90 days mortality.

## Data Availability

Data can be given on demand

## References

1. Couinaud C. Surgical anatomy of the liver revisited. Paris, France: Self-printed; 1989. □

2. Yamamoto M, Ariizumi SI. Glissonean pedicle approach in liver surgery. Ann Gastroenterol Surg. 2018 Feb 13;2(2):124–128.

3. Takasaki K. Glissonean pedicle transection method for hepatic resection: a new concept of liver segmentation. J Hepatobiliary Pancreat Surg. 1998;5:286–291. □

4. Tung TT, Quang ND. A new technique for operating on the liver. Lancet. 1963;26:192–193.

5. Lin TY, Chem KM, Liu TK. Total right hepatic lobectomy for primary hepatoma. Surgery. 1960;48:1048–1060. □

6. Robert J. Aragon, Naveenraj L. Solomon Techniques of hepatic resection. J Gastrointest Oncol 2012;3:28–40.

7. Blumgart LH, Belghiti J. Surgery of the liver, biliary tract, and pancreas. 4th ed. Philadelphia, PA: Saunders Elsevier, 2007.

8. Gurusamy KS, Pamecha V, Sharma D, Davidson BR. Techniques for liver parenchymal transection in liver resection. Cochrane Database Syst Rev. 2009 Jan 21;(1):CD006880.

9. Takasaki K. Glissonean pedicle transection method for hepatic resection: a new concept of liver segmentation. J Hepatobiliary Pancreat Surg. 1998;5(3):286–91.

10. Lee N, Cho CW, Kim JM, Choi GS, Kwon CHD, Joh JW. Application of temporary inflow control of the Glissonean pedicle method provides a safe and easy technique for totally laparoscopic hemihepatectomy by Glissonean approach. Ann Surg Treat Res. 2017 May;92(5):383–386.

11. Kim SH, Kim KH, Kirchner VA, Lee SK. Pure laparoscopic right hepatectomy for giant hemangioma using anterior approach. Surg Endosc. 2017 May;31(5):2338–2339.

12. Cho W, Kwon CHD, Choi JY, Lee SH, Kim JM, Choi GS et al. Impact of technical innovation on surgical outcome of laparoscopic major liver resection: 10 years’ experience at a large-volume center. Ann Surg Treat Res. 2019 Jan;96(1):14–18.

13. Launois B, Maddern G, Tay KH. The Glissonian approach of the hilum. Swiss Surg. 1999;5(3):143–6.

14. Virani S, Michaelson JS, Hutter MM, Lancaster RT, Warshaw AL, Henderson WG, Morbidity and mortality after liver resection: results of the patient safety in surgery study. Am Coll Surg. 2007 Jun;204(6):1284–92.

15. Dokmak S, Ftériche FS, Borscheid R, Cauchy F, Farges O, Belghiti J. 2012 Liver resections in the 21st century: we are far from zero mortality. HPB (Oxford). 2013;15(11):908–15.

16. Martínez-Mier G, Esquivel-Torres S, Alvarado-Arenas RA, Ortiz-Bayliss AB, Lajud-Barquín FA, Zilli-Hernandez S. Liver resection morbidity, mortality, and risk factors at the departments of hepatobiliary surgery in Veracruz, Mexico. Rev Gastroenterol Mex. 2016 Oct-Dec;81(4):195–201

17. Gilg S, Sparrelid E, Isaksson B, Lundell L, Nowak G, Strömberg C. Mortality-related risk factors and long-term survival after 4460 liver resections in Sweden-a population-based study. Langenbecks Arch Surg. 2017 Feb;402(1):105–113.

